# Role of Cardiorespiratory Fitness in Modulating Cardiotoxicity in Long-Term Cancer Survivors Exposed to Anthracycline Therapy

**DOI:** 10.1101/2025.03.28.25324862

**Authors:** Olga H. Toro-Salazar, Linda S. Pescatello, May Ling Mah, Caroline Wilhelm, Andrea D. Orsey, Tiffany Berthod, Maua H. Mosha, Michael Brimacombe, Corbinian Wanner, Michelle Slawiniski, Kan N. Hor

## Abstract

**Background:** Childhood cancer survivors (CCS) often experience cardiotoxicity due to cancer treatment. Furthermore, low cardiorespiratory fitness (CRF) is a strong predictor of mortality.

**Objective:** This study investigates the effects of a 16-week supervised aerobic exercise intervention on CRF changes, as measured by VO_2peak_ and parameters of cardiac structure and function via cardiac magnetic resonance imaging (CMR).

**Methods:** CCS were enrolled >2 years post-treatment and underwent cardiotoxicity risk stratification, cardiopulmonary stress test to determine CRF, and CMR before and after exercise intervention. Subclinical myocardial dysfunction was assessed by segmental strain abnormality, with specific focus on the number of segments showing peak circumferential strain magnitude (εcc) ≤10% and εcc ≤17%.

**Results:** Forty-seven subjects (10-25 years, median 22.0 years, IQR 9.0), 57.4% male, and 76% white were enrolled. Thirty-one patients completed the exercise intervention. VO_2peak_ increased ≥ 1 mL/kg/min in 16/31 (51.6%) of subjects. These subjects were labeled “responders”, demonstrated a median increase in VO_2peak_ of 3.8 mL/kg/min. Responders showed significant increase in left (p=0.0348) and right (p=0.0390) ventricular end diastolic volume and stroke volume index (p=0.0141). Left ventricular ejection fraction continued to decline in non-responders with a net difference of 2.45% favoring responders (p=0.007). At baseline, 11 subjects met high-risk definition based on segmental strain abnormalities (i.e. ≥2 segments with peak circumferential strain magnitude (εcc) ≤10% or ≥9 segments ≤17%). High risk subjects showed significant improvement in εcc with a post-intervention reduction in number of segments ≤17% from 8.9 to 5.9 (p=0.014) and ≤10% from 1.8 to 0.3 (p=0.022).

**Conclusions:** CCS demonstrated increase in post-intervention VO_2peak_ and improvement in CMR parameters of structure and function. Those classified as high-risk based on εcc abnormalities benefited the most.

**Trial registration:** NCT, NCT04036032. Unique Protocol ID: 14-110. Registered 25 July 2019 - Retrospectively registered, https://classic.clinicaltrials.gov/ct2/show/NCT04036032

## Background

Although advances in pediatric cancer treatment have raised survival rates to over 80%, there is an increased morbidity and mortality in childhood cancer survivors (CCS) due to the toxic side effects of cancer therapy.^1–3^ Among CCS in the United States, over 50% are exposed to cardiotoxic treatments that increase their risk of heart failure. Cardiovascular disease (CVD) is the second leading cause of morbidity and mortality in CCS after recurrent malignancy, with a 5-year survival rate of less than 50% after developing clinical heart failure.^1,2,4,5^

Cardiorespiratory fitness (CRF) integrates the cardiovascular, respiratory, and musculoskeletal systems and reflects overall health.^4^ CRF declines during and after cancer treatment due to treatment-related deconditioning and adverse cardiovascular and musculoskeletal side effects. Low CRF strongly predicts mortality in both healthy individuals and those with CVD.^5–7^ Exercise is recommended for preventing and managing many chronic diseases to reduce the risk of premature mortality.^8–10^ Regular physical activity may reduce the risk of cancer relapse and mitigate the adverse effects of treatment. ^11,12,13^

Peak oxygen consumption (VO_2peak_) is the gold standard assessment of CRF.^13,14^ Studies conducted in children and adults have demonstrated that improvements in VO_2peak_ ≥1.0 mL/kg/min are clinically significant. ^6,15,16^ VO_2peak_ serves as a health indicator for cardio-metabolic health, premature CVD, and all-cause, cardiac, and cancer-related mortality. In adults, an increase in VO_2peak_ ≥1.0 mL/kg/min decreases the incidence of all-cause, cardiac, and cancer-related mortality over a median follow-up of 4.5–7.9 years by 9–10%, 15%, and 16%, respectively.^6,8,15^ Exercise training impact on left ventricular (LV) structure includes LV hypertrophy, cardiac chamber enlargement, and increased stroke volume, the hallmarks of endurance-trained athletes.^17^ Aerobic exercise therapy also modulates gene expression pathways that may promote cardiac remodeling.^18^ Cardiac magnetic resonance (CMR) imaging is utilized for assessment of LV structure and global and regional myocardial function, and is able to detect early cardiac injury and adverse cardiac remodeling as a result of cancer treatment.^19,20^ CMR myocardial tagging is the reference modality for evaluating myocardial strain.^21^ Measures of myocardial strain by tagged cine imaging and strain encoded CMR (SENC) are increasingly being used to detect subclinical myocardial dysfunction in cancer therapy-related cardiac dysfunction and other non-ischemic cardiomyopathies.^22^

The purpose of this study was to examine the effect of a 16-week supervised aerobic exercise intervention in CCS at the subject’s local YMCA on changes in CRF and parameters of cardiac structure and function assessed by VO_2peak_ measurements and CMR.

## Methods

This study was a prospective multicenter pilot study. This study was approved by the Institutional Review Board at Connecticut Children’s and Nationwide Children’s Hospital. A standard of practice approach was utilized in the study design with use of predicted values as normative data for the cardio-pulmonary stress test (CPET) and Z-scores for CMR parameters.

The study flow diagram is shown in **Figure 1**. Eligible subjects were identified from a cardio-oncology registry between June 2015 and March 2019. Included were CCS over 9 years old exposed to anthracyclines. Exclusions were pregnancy, contraindications for CMR, exercise, or CPET (per the American College of Sports Medicine). Chart reviews were conducted to identify risk factors of cardiotoxicity.^1,2,23,24^ A cardiotoxicity risk profile and a raw score was assigned to classify subjects as low (0-5), moderate (6-10), or high risk (≥11), **Figure 2**.

**Figure 1.**
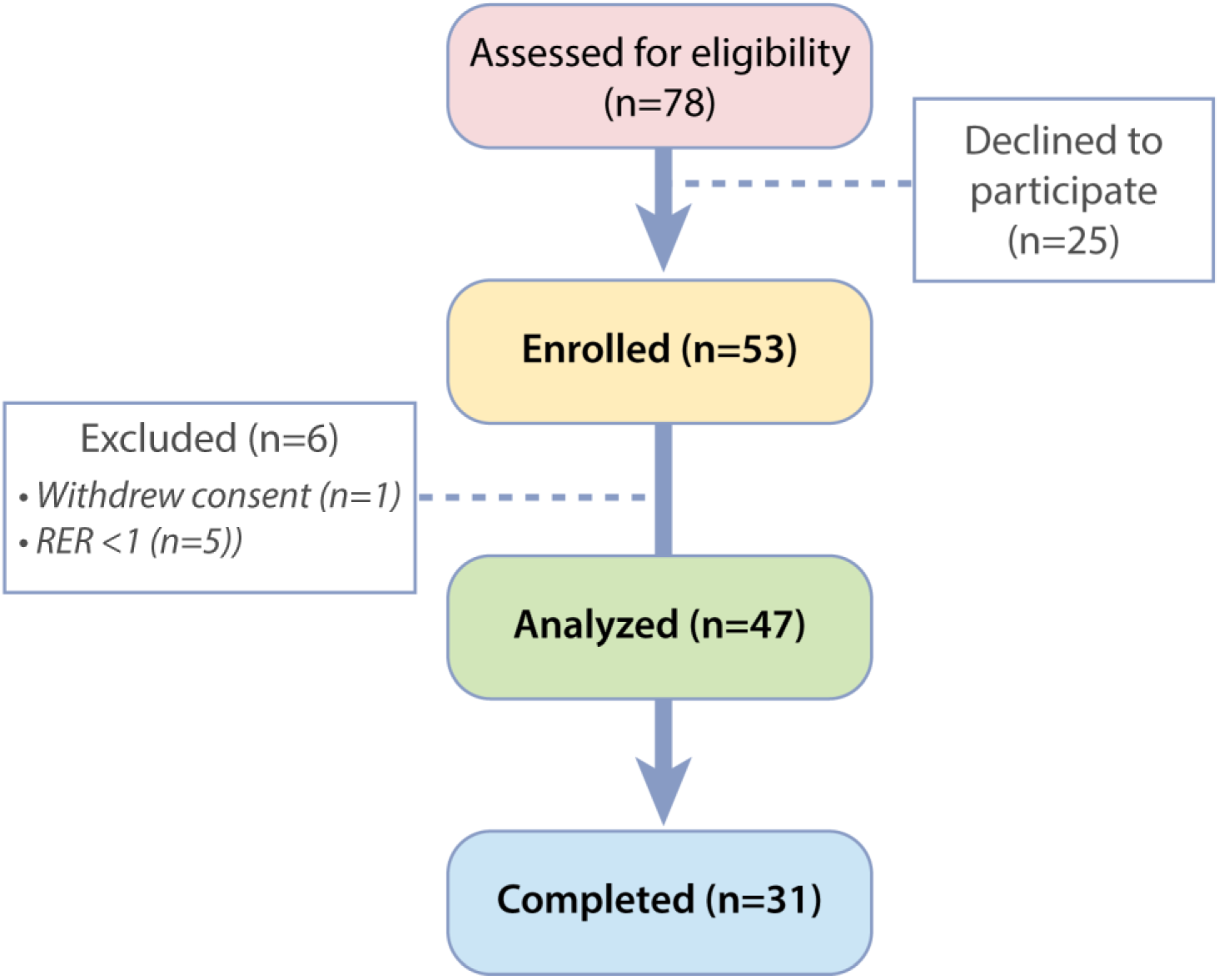
Study Flow Overview:

**Figure 2.**
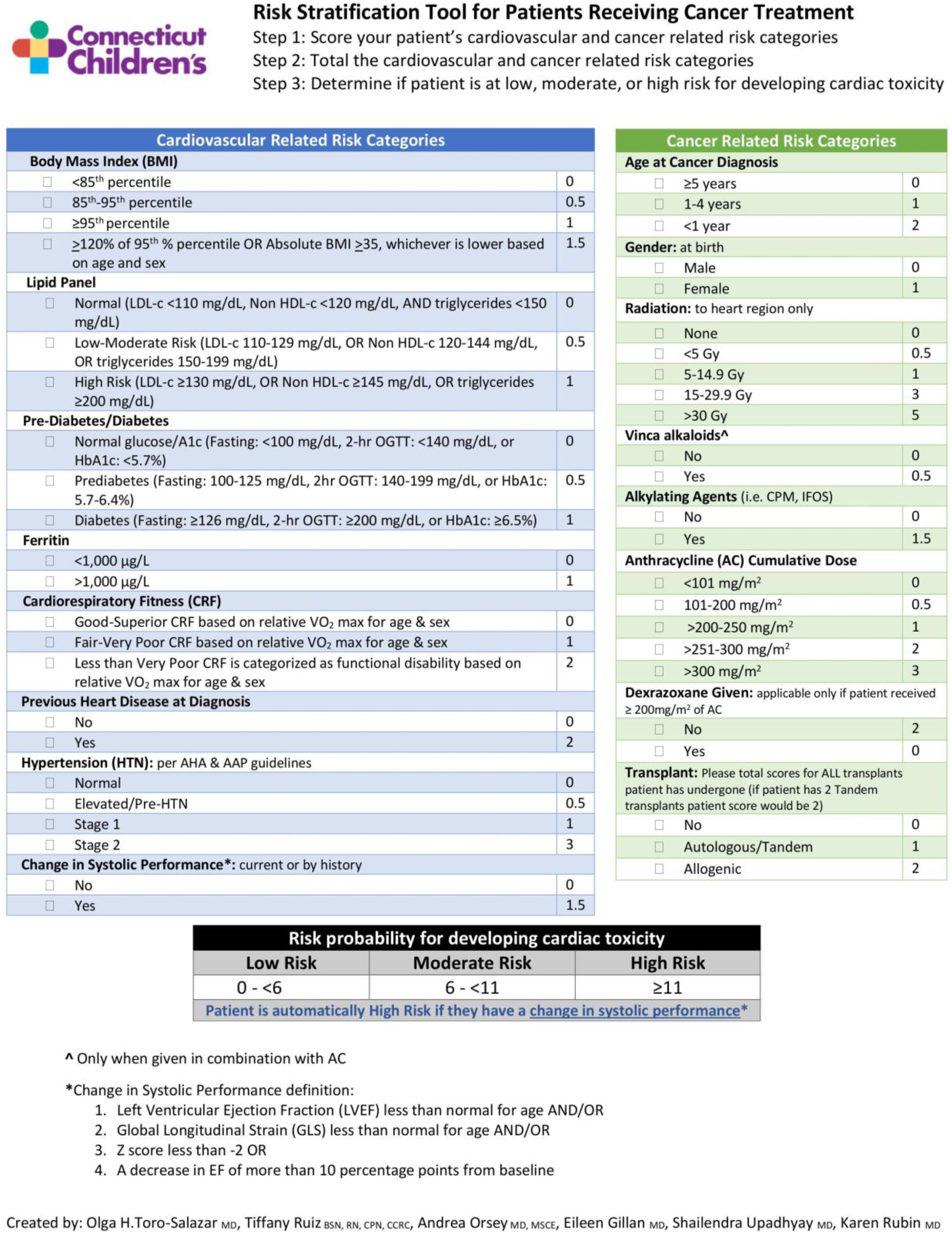
Risk Stratification Scoring Pathway. *Our team created a cardio-oncology registry to document the diagnosis, treatment, clinical course, and outcomes of children exposed to cardiotoxic therapy, aiding in the development of comprehensive risk models for cardiotoxicity. Factors associated with increased mortality risk included longer post-chemotherapy intervals, younger age at diagnosis, total cumulative anthracycline dose, history of bone marrow transplant, and a solid tumor diagnosis.*^49^ *Based on these results, literature review, and traditional cardiovascular disease risk factors, we developed a composite cardiotoxicity scoring system to classify patients as low, moderate, and high risk. This scoring system has been validated and significantly correlates with LV-EF (R: −0.6307, p<.0001)*.

Subjects were evaluated before and after the YMCA exercise intervention. Participants were categorized as responders (change in VO_2peak_ ≥1.0 mL/kg/min) and non-responders (change in VO_2peak_ ≤1.0 mL/kg/min). VO_2peak_ was defined as the mean value measured within the last 20 seconds of the exercise test, expressed in mL/kg/min and L/min. Predicted VO_2peak_ was calculated using the FRIEND registry equation, based on age, gender, and anthropometric data.^25^ Changes in CMR parameters measured included LV and right ventricular (RV) volume, mass, global function, peak global and segmental circumferential strain (εcc), and longitudinal strain (ειι) magnitude. Methods for the exercise intervention, CPET, CMR protocol, and secondary endpoints are in the Supplemental Material.

### Exercise Intervention

The YMCA was selected because it offered a free “Living Healthy, Living Strong” class and personal trainers who specialized in working with patients with cancer. At baseline, a personal trainer met with each CCS to provide a customized exercise program based on the subject’s Toolbox exercise modality of choice.^26^ The Toolbox of exercise modalities (e.g., cycling, rowing, stair climbing, jogging, swimming, etc.) consisted of a carefully selected set of physical activities of similar aerobic intensity designed by a personal trainer to meet the specific needs and goals of the subject during training.^27^ As part of the baseline assessment (week 0), all participants were instructed to maintain their baseline physical activity level and were given an educational brochure regarding the benefits of exercise. At week 1, participants were asked to exercise for 15 minutes per day progressing to 45 minutes per day, five days/week or as tolerated by the end of week 4. The goal during each session was for the subject to exercise at 50-80% of their age-adjusted maximum heart rate as determined by the CPET. The exercise intervention continued for an additional 12 weeks (ending at 16 weeks after study entry), and participants were asked to meet with their personal trainer every 4 weeks (±1 week) for a total of 5 visits over the course of the study. Each session was completed independently by those 18 years or older, or under the guidance and supervision of parent/legal guardian for those younger than 18 years, and scheduled at their convenience. Either the personal trainer or study staff conducted weekly coaching phone calls to provide encouragement and address any barriers to exercise performance.

All subjects received a Fitbit tracker (Fitbit-Alta HR™, Fitbit, Inc. San Francisco, CA) to collect actigraphy data, including total step counts. During the 16-week exercise intervention, data collection tracked changes in physical activity (total step counts) over time as classified into fairly active (moderate) or very active (intense) physical activity. Adherence to the exercise intervention was calculated using participants Fitbit profiles from the prescribed vs. completed number of exercise sessions.

### Statistical Analysis

Descriptive statistics were performed on subject demographics, CMR, and CPET parameters. Results are expressed as median and inter-quartile range (IQR) unless otherwise indicated. Pearson’s correlation tested the linear relationships among VO_2peak_, CMR parameters, and cardiotoxicity risk factor scores. Pearson correlation was used where the responses analyzed were continuous in nature. Differences between proportions were assessed using chi-square or Fisher exact tests. Multiple regression tested the relationships among VO_2peak_ and CMR parameters after adjusting for cardiotoxicity risk scores. Differences between medians were assessed using Mann-Whitney test. Tables reflect means and medians to demonstrate the stability of the data. In Table 5, medians were included since non-parametric, rank-based p-values were generated due to usage of rank-based Mann-Whitney test. Statistical analyses were conducted using SAS 9.4 (SAS Institute Inc., Cary, NC) with significance level set at p <0.05.

## Results

### Study Population

Fifty-three of 78 eligible subjects enrolled in the study. The main barriers to participation were access to a local YMCA, time commitment, and involvement in other sport/exercise programs. Demographic and clinical characteristics of study participants are shown in **Table 1**. Of 53 consented subjects, 47 continued to the exercise intervention stage as five subjects were unable to complete the baseline CPET and one subject withdrew their consent (**Figure 1**). The 47 participants were 57.4% male and had a median age of 22.0 years (IQR 9.0 years), with a range of 10-25 years. The most common cancer diagnoses were lymphoma (29.8%), acute lymphoblastic leukemia (27.7%), sarcoma (17%), and acute myelogenous leukemia (8.5%).

**Table 1.** Demographics.

| Variables | All Participants | Completed Study | Dropouts | P-value comparing Completed and Dropouts |
| --- | --- | --- | --- | --- |
| **Gender; n (%)** |  |  |  |  |
| Male | 27 (57.4) | 18 (58.1) | 10 (62.5) | 0.76 |
| Female | 20 (42.6) | 13 (41.9) | 6 (37.5) |  |
| **Race; n (%)** |  |  |  |  |
| White | 35 (74.5) | 26 (83.9) | 9 (56.3) | 0.080 |
| Non-white | 12 (25.5) | 5 (16.1) | 7 (43.7) |  |
| **Ethnicity; n (%)** |  |  |  |  |
| Hispanic | 5 (10.6) | 1 (3.2) | 4 (25) | 0.040 |
| Non-Hispanic | 42 (89.4) | 30 (96.8) | 12 (75) |  |
| **Demographic Factors; median (IQR)** |  |  |  |  |
| Age at Baseline | 22.0 (9.0) | 24.0 (10.0) | 22.0 (7.8) | 0.96 |
| Time from Last Chemotherapy Treatment (years) | 7.4 (7.0) | 7.8 (8.7) | 7.1 (7.0) | 0.57 |
| Height, cm | 169.0 (16.0) | 170.0 (21.3) | 167.3 (12.3) | 0.98 |
| Weight, kg | 76.5 (30.1) | 68.1 (29.1) | 83.2 (18.9) | 0.080 |
| BSA, m<sup>2</sup> | 1.9 (0.5) | 1.8 (0.5) | 2.0 (0.2) | 0.21 |
| BMI, kg/m<sup>2</sup> | 24.6 (8.7) | 24.0 (6.7) | 29.8 (9.6) | 0.040 |
| BP systolic (n = 46) | 113.5 (17.5) | 110 (17.5) | 118.5 (20.0) | 0.45 |
| BP diastolic (n = 46) | 68.5 (17.0) | 68.5 (6.25) | 69.0 (20.3) | 0.34 |
| Cardiotoxicity risk score | 7.0 (4.5) | 7.0 (6.0) | 7.0 (3.4) | 0.97 |
| **Diagnosis; n (%)** |  |  |  |  |
| ALL* | 13 (27.7) | 9 (29) | 4 (25) | -- |
| AML | 4 (8.5) | 4 (12.9) | 0 | -- |
| Lymphoma* | 14 (29.8) | 7 (22.6) | 7 (43.8) | -- |
| Other* | 16 (34.0) | 11 (35.5) | 5 (31.2) | -- |
| **Radiation to the Chest; n (%)** |  |  |  |  |
| Yes | 15 (31.9) | 9 (29) | 6 (37.5%) | 0.74 |
| No | 32 (68.1) | 22 (71) | 10 (62.5%) | -- |

\*Subjects with secondary malignancy
**Abbreviations:** Body Surface Area (BSA), Body Mass Index (BMI), Blood Pressure (BP), Acute Lymphocytic Leukemia (ALL), Acute Myeloid Leukemia (AML)

Thirty-one subjects completed the exercise intervention. A higher proportion of those who completed the exercise intervention were white (83.9%) and had a significantly lower median BMI (24.0 kg/m² versus 29.8 kg/m²; p=0.040) (**Table 1**).

### Cardiotoxicity Risk Score

Subjects received a composite risk score based on cardiotoxicity and traditional CVD risk factors (**Figure 2**). The median score was 7 (IQR 4.5) (**Table 1**). At baseline, the risk score was inversely related to LV stroke volume index (SVi) (R: −0.2986, p=0.042), LV ejection fraction (EF) (R: −0.4403, p=0.002), RV-SVi (R: −0.2988, p=0.041), global peak longitudinal strain (ειι) (R: 0.3636, p=0.013), and positively correlated with LV end-systolic fiber stress (ESFS) (R: 0.4486, p=0.002) and decreased peak circumferential strain magnitude (εcc) ≤17% (R: 0.36, p=0.013) (**Table 2**).

**Table 2.** Correlation of VO_2peak_ and Cardiotoxicity Risk score with BMI and CMR Parameters at Baseline.

|  | **Coefficient** | **p** |
| --- | --- | --- |
| **VO<sub>2peak</sub>, mL/kg/min** |  |  |
| **O<sub>2</sub> pulse, mL/beat** | 0.3497 | 0.016 |
| **BMI, kg/m<sup>2</sup>** | -0.6968 | <0.001 |
| **BSA, m<sup>2</sup>** | -0.4944 | <0.001 |
| **LV-EDVi, mL/m<sup>2</sup>** | 0.3021 | 0.039 |
| **LV-EDV z-score** | 0.3106 | 0.034 |
| **LV-SVi, mL/m<sup>2</sup>** | 0.4226 | 0.003 |
| **LV Mass index, g/m<sup>2</sup>** | 0.4538 | 0.001 |
| **RV-EDV z-score** | 0.3012 | 0.042 |
| **RV-EDVi, mL/m<sup>2</sup>** | 0.3855 | 0.007 |
| **RV-ESVi, mL/m<sup>2</sup>** | 0.3056 | 0.037 |
| **RV-SVi, mL/m<sup>2</sup>** | 0.4214 | 0.003 |
| **Cardiotoxicity risk score** |  |  |
| **LV-SVi, mL/m<sup>2</sup>** | -0.2986 | 0.042 |
| **LV-EF, %** | -0.4403 | 0.002 |
| **LV-EF z-score** | -0.4360 | 0.002 |
| **LV-ESFS, (g/cm<sup>2</sup>** | 0.4486 | 0.002 |
| **LV-ESFS z-score** | 0.4413 | 0.002 |
| **RV-EDV z-score** | -0.3570 | 0.015 |
| **RV-SVi, mL/m<sup>2</sup>** | -0.2988 | 0.041 |
| **ε<sub>u</sub>, %** | 0.3636 | 0.013 |

**Abbreviations:** Body Mass Index (BMI), Body Surface Area (BSA), Left Ventricle (LV), End Diastolic Volume (EDV), Stroke Volume Index (SVi), Right Ventricle (RV), Ejection Fraction (EF), End Systolic Fiber Stress (ESFS), Peak Longitudinal Strain Magnitude (ε<sub>u</sub>).

### Baseline Cardiorespiratory Fitness

Baseline CPET parameters for all 31 subjects are shown in **Table 3**. Participants had low CRF, with a median indexed VO_2peak_ of 32.9 mL/kg/min (IQR 13.4), predicted 41.7 mL/kg/min (IQR 9.4). Functional disability (VO_2peak_ ≤18 mL/kg/min) was present in 4 subjects. Baseline VO_2peak_ was inversely correlated with BMI (p<0.001) and positively correlated with volumetric and functional CMR parameters (**Table 2).**

**Table 3.** Subject Baseline Cardiorespiratory Parameters.

| | Mean ± SD | Median (IQR) |
| --- | --- | --- |
| VO<sub>2</sub> predicted, mL/kg/min | 43.1 ± 6.6 | 41.7 (9.4) |
| VO<sub>2</sub>, mL/kg/min | 33.7 ± 9.3 | 32.9 (13.4) |
| Anaerobic threshold, L/min | 1.7 ± 0.6 | 1.7 (0.6) |
| Anaerobic threshold, % predicted peak VO<sub>2</sub> | 56.6 ± 17.1 | 51 (15) |
| O<sub>2</sub> pulse, ml/beat | 13.2 ± 3.4 | 13.3 (5.3) |
| RER, peak | 1.2 ± 0.1 | 1.2 (0.1) |
| METS | 10.0 ± 2.7 | 10 (3.4) |

*Abbreviations: Respiratory Exchange Rate (RER) Metabolic Equivalents (METS).*

### VO_2peak_ Response to the Exercise Intervention

Subjects with a positive change in VO_2peak_ ≥1 mL/kg/min (15 participants, 48.4%) were classified as responders, and those with a negative response (16 participants) as non-responders (**Figure 3**). Baseline and post-intervention CPET variables among responders and non-responders are described in **Table 4**. Responders demonstrated significant increases in CRF (median change of VO_2peak_: +3.8 mL/kg/min) whereas VO_2peak_ continued to decline in non-responders (median change: −1.85 mL/kg/min), with a 5.65 mL/kg/min net difference favoring responders to the exercise intervention (p<0.001). Similarly, O_2_ pulse peak increased in responders (median change: +1.2 mL/beat) and declined in non-responders (median change: − 1.15 mL/beat), with a net difference of 2.35 mL/beat favoring responders (p <0.001) **(Table 4).** Change in VO_2peak_ was inversely correlated with change in BMI (R: −0.38, p=0.036). There were no exercise-related serious adverse events.

**Figure 3.**
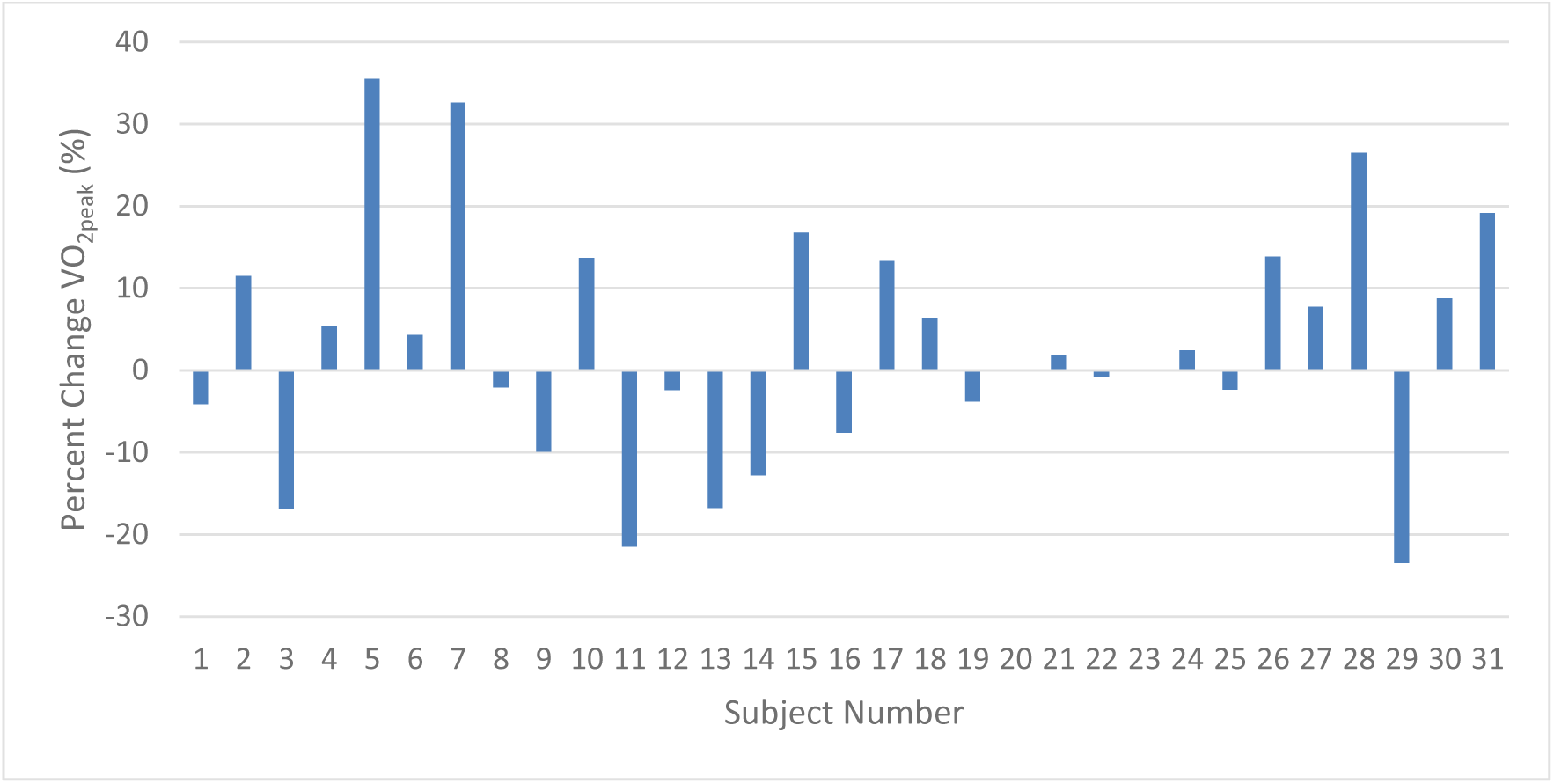
Change in Indexed VO_2peak_ after Intervention. *Percent change in indexed VO_2peak_, measured in mL/kg/min, in all study participants pre- and post-intervention. 15 individuals demonstrated an improvement of ≥1ml/kg/min in VO_2peak_ and were classified as responders*.

**Table 4.** Baseline and Follow-up Cardiorespiratory Parameters in Responders and Non-Responders.

| Outcome | Responders |  |  | Non-Responders |  |  | Mann-Whitney P-value |
| --- | --- | --- | --- | --- | --- | --- | --- |
|  | n | Median | IQR | n | Median | IQR |  |
| VO<sub>2</sub>peak, mL/kg/min |  |  |  |  |  |  |  |
| Baseline | 15 | 33.1 | 16.0 | 16 | 38.0 | 15.5 | 0.37 |
| Post-Intervention | 15 | 38.5 | 15.2 | 16 | 32.1 | 14.1 | 0.21 |
| Δ Post-Intervention | 15 | 3.8 | 4.5 | 16 | -1.85 | 4.85 | <0.001 |
| VO<sub>2</sub>peak, % predicted |  |  |  |  |  |  |  |
| Baseline | 15 | 46.3 | 13.9 | 16 | 41.7 | 6.63 | 0.38 |
| Post-Intervention | 15 | 46.3 | 13.8 | 16 | 41.0 | 6.63 | 0.59 |
| Δ Post-Intervention | 15 | 0.0 | 0.3 | 16 | 0.0 | 0.40 | 0.59 |
| O<sub>2</sub> pulse peak, mL/beat |  |  |  |  |  |  |  |
| Baseline | 13 | 13.2 | 7.9 | 14 | 13.45 | 4.75 | 0.37 |
| Post-Intervention | 13 | 14.6 | 10.6 | 14 | 11.90 | 5.58 | 0.18 |
| Δ Post-Intervention | 13 | 1.2 | 2.45 | 14 | -1.15 | 1.83 | <0.001 |
| Heart rate peak |  |  |  |  |  |  |  |
| Baseline | 14 | 68.0 | 21.25 | 16 | 64.0 | 11.25 | 0.46 |
| Post-Intervention | 15 | 69.0 | 19.0 | 16 | 67.0 | 12.25 | 0.59 |
| Δ Post-Intervention | 14 | 1.0 | 5.0 | 16 | 0.0 | 12.25 | 0.73 |
| RER |  |  |  |  |  |  |  |
| Baseline | 15 | 1.19 | 0.13 | 16 | 1.17 | 0.11 | 0.59 |
| Post-Intervention | 13 | 1.15 | 0.08 | 14 | 1.19 | 0.12 | 0.31 |
| Δ Post-Intervention | 13 | -0.03 | 0.09 | 14 | 0.03 | 0.12 | 0.080 |
| Average VE peak Value, L/min |  |  |  |  |  |  |  |
| Baseline | 15 | 98.8 | 49.2 | 16 | 92.85 | 41.82 | 0.59 |
| Post-Intervention | 15 | 104.2 | 47.1 | 16 | 94.15 | 45.02 | 0.59 |
| Δ Post-Intervention | 15 | 6.0 | 13.1 | 16 | -2.55 | 19.32 | 0.049 |
| SBP peak, mmHg |  |  |  |  |  |  |  |
| Baseline | 15 | 110.0 | 15.7 | 16 | 110.0 | 18.75 | 0.43 |
| Post-Intervention | 15 | 110.0 | 23.0 | 16 | 109.5 | 17.25 | 0.89 |
| $\Delta$ Post-Intervention | 15 | 1.5 | 14.2 | 16 | -2.0 | 20.25 | 0.14 |
| **DBP peak, mmHg** |  |  |  |  |  |  |  |
| Baseline | 14 | 65.5 | 24.2 | 16 | 68.5 | 16.0 | 1.0 |
| Post-Intervention | 15 | 64.0 | 18.0 | 16 | 61.0 | 18.5 | 0.59 |
| $\Delta$ Post-Intervention | 14 | -2.0 | 15.0 | 16 | -1.0 | 14.5 | 1.0 |

**Abbreviations:** Respiratory Exchange Rate (RER) Minute Ventilation (VE), Systolic Blood Pressure (SBP), Diastolic Blood Pressure (DBP).

**Table 5.** Baseline and Follow-up CMR Parameters in Responders and Non Responders.

| Outcome | Responders |  |  | Non-Responders |  |  | Mann-Whitney<br>P-value |
| --- | --- | --- | --- | --- | --- | --- | --- |
|  | n | Median | IQR | n | Median | IQR |  |
| LV-EDVi, mL/m<sup>2</sup> |  |  |  |  |  |  |  |
| Baseline | 15 | 81.0 | 28.12 | 16 | 80.81 | 15.55 | 0.85 |
| Post-Intervention | 15 | 85.48 | 26.93 | 16 | 77.59 | 18.67 | 0.59 |
| Δ Post-Intervention | 15 | 2.57 | 5.58 | 16 | -0.88 | 8.87 | 0.59 |
| LV-ESVi, mL/m<sup>2</sup> |  |  |  |  |  |  |  |
| Baseline | 15 | 36.0 | 11.86 | 16 | 32.32 | 7.46 | 0.59 |
| Post-Intervention | 15 | 35.44 | 10.58 | 16 | 34.06 | 7.38 | 0.59 |
| Δ Post-Intervention | 15 | 0.0 | 3.48 | 16 | 1.49 | 4.32 | 0.37 |
| LV-SVi, mL/m<sup>2</sup> |  |  |  |  |  |  |  |
| Baseline | 15 | 45.5 | 16.24 | 16 | 44.31 | 7.91 | 0.59 |
| Post-Intervention | 15 | 45.88 | 15.91 | 16 | 45.21 | 10.77 | 0.85 |
| Δ Post-Intervention | 15 | 1.76 | 5.63 | 16 | -2.06 | 5.13 | 0.38 |
| LV-EF, % |  |  |  |  |  |  |  |
| Baseline | 15 | 55.6 | 3.4 | 16 | 57.8 | 7.38 | 0.10 |
| Post-Intervention | 15 | 56.3 | 5.2 | 16 | 56.5 | 4.6 | 0.85 |
| Δ Post-Intervention | 15 | 0.30 | 3.1 | 16 | -2.15 | 3.53 | 0.007 |
| LV mass index, g/m<sup>2</sup> |  |  |  |  |  |  |  |
| Baseline | 15 | 57.93 | 21.0 | 16 | 56.0 | 9.08 | 0.59 |
| Post-Intervention | 15 | 54.75 | 23.44 | 16 | 48.63 | 11.81 | 0.21 |
| Δ Post-Intervention | 15 | -3.93 | 10.83 | 16 | -3.21 | 8.68 | 0.85 |
| LV mass/volume, g/mL |  |  |  |  |  |  |  |
| Baseline | 15 | 0.65 | 0.15 | 16 | 0.70 | 0.14 | 0.85 |
| Post-Intervention | 15 | 0.70 | 0.23 | 16 | 0.65 | 0.12 | 0.11 |
| Δ Post-Intervention | 15 | -0.01 | 0.16 | 16 | -0.025 | 0.105 | 0.59 |
| RV-EDVi, mL/m<sup>2</sup> |  |  |  |  |  |  |  |
| Baseline | 15 | 75.91 | 40.92 | 16 | 84.52 | 23.19 | 0.37 |
| Post-Intervention | 15 | 84.0 | 33.44 | 16 | 80.95 | 21.12 | 0.59 |
| Δ Post-Intervention | 15 | 3.10 | 11.59 | 16 | -1.89 | 11.29 | 0.049 |
| RV-ESVi, mL/m<sup>2</sup> |  |  |  |  |  |  |  |
| Baseline | 15 | 34.0 | 16.03 | 16 | 37.07 | 11.13 | 0.37 |
| Post-Intervention | 15 | 33.28 | 16.16 | 16 | 34.38 | 13.36 | 0.85 |
| Δ Post-Intervention | 15 | 1.46 | 4.07 | 16 | -1.07 | 4.98 | 0.21 |
| RV-SVi, mL/m<sup>2</sup> |  |  |  |  |  |  |  |
| Baseline | 15 | 45.0 | 17.99 | 16 | 44.97 | 8.85 | 0.85 |
| Post-Intervention | 15 | 45.88 | 15.59 | 16 | 45.30 | 8.43 | 0.59 |
| $\Delta$ Post-Intervention | 15 | 2.61 | 6.75 | 16 | -1.26 | 6.04 | 0.049 |
| **RV-EF, %** |  |  |  |  |  |  |  |
| Baseline | 14 | 57.65 | 6.4 | 16 | 56.2 | 7.8 | 1.0 |
| Post-Intervention | 15 | 56.1 | 4.5 | 15 | 57.6 | 5.5 | 0.27 |
| $\Delta$ Post-Intervention | 14 | -0.65 | 4.33 | 15 | -1.5 | 5.1 | 0.86 |

**Abbreviations:** *Left Ventricle (LV), End Diastolic Volume Index (EDVi), End Systolic Volume*
*Index (ESVi), Stroke Volume Index (SVi), Ejection Fraction (EF), Right Ventricle (RV).*

### Baseline CMR Parameters

Seventeen of 47 participants (36.2%) had an LV-EF <55%, 14 (29.8%) an LV-EF of 50-54%, and 3 (6.4%) LV-EF <50%. Global RV systolic dysfunction (RV-EF <49%) was present in 1 participant (2.1%). Decreased εcc magnitude ≤17% was found in 24 participants (51.1%), and decreased ειι ≤10% in 27 participants (57.5%) (**Supplemental Table 1**). Decreased ειι correlated with cumulative anthracycline dose (R: 0.35, p<0.020). At baseline, 11 subjects met high-risk definition based on segmental strain abnormalities (i.e. ≥2 segments with peak circumferential strain magnitude (εcc) ≤10% or ≥9 segments ≤17%) **Figure 4**). Additional baseline CMR parameters are included in **Supplemental Table 1**.

**Figure 4.**
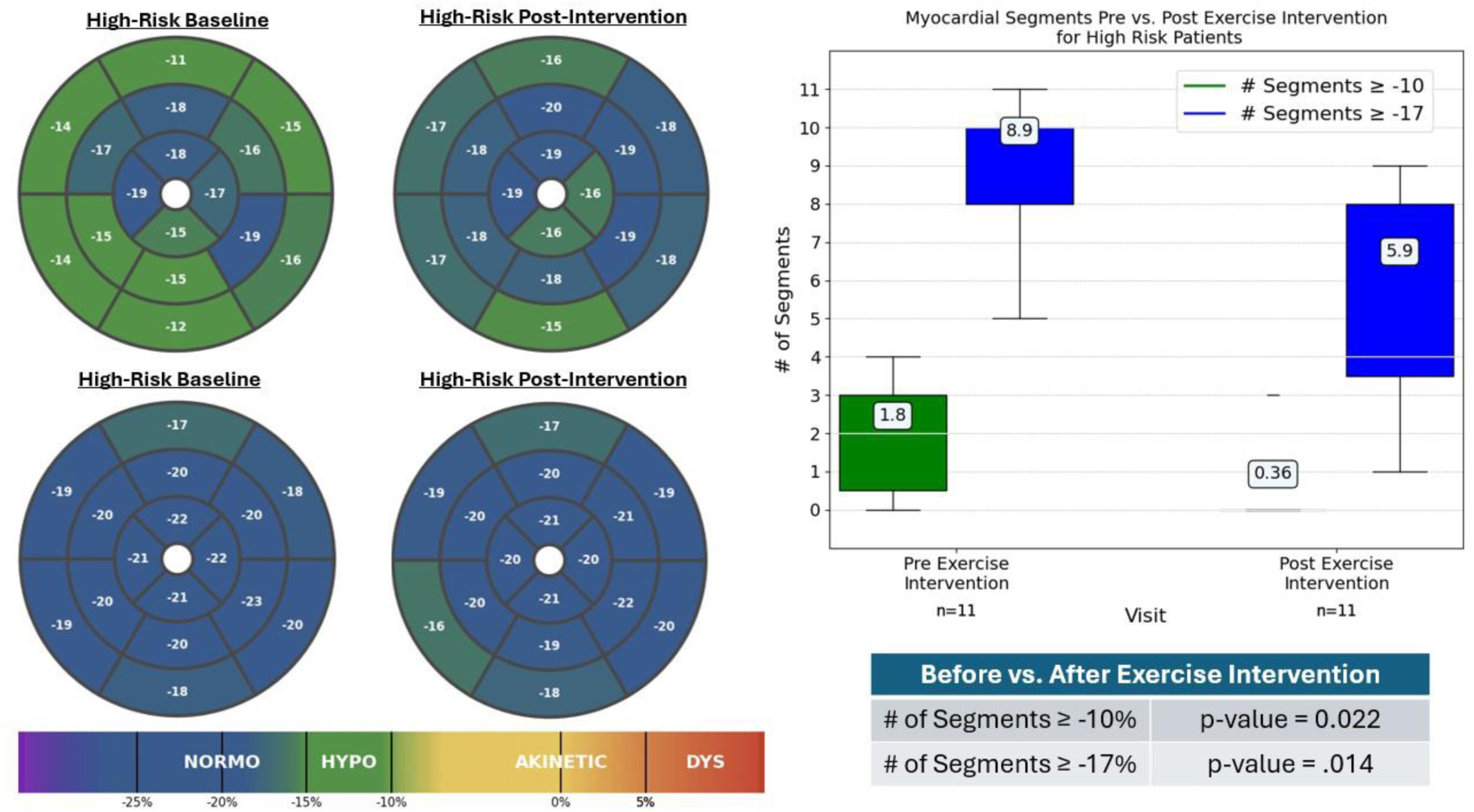
Segmental Circumferential Strain Abnormalities Before and After Exercise Intervention. ***(A)*** *Polar map shows segmental circumferential strain from cine tagged acquisition at the basal, mid, and apical levels of the short-axis view at baseline and after the exercise intervention. The color scale indicates peak strain magnitude in the myocardium. Subjects with 2 or more segments ≤10% or 9 or more segments ≤17% were classified as high risk. **(B)** Change in number of abnormal myocardial segments in the high-risk group before and after the exercise intervention*.

### CMR Parameters Response to Exercise Intervention

Imaging parameters of LV and RV global and regional myocardial function among responders and non-responders are included in **Table 5 and Figure 5**. Change in VO_2peak_ inversely correlated with change in BMI and positively correlated with change in LV end diastolic volume index (EDVi) (p=0.0348), RV-EDVi (p=0.0390), and LV-SVi (p=0.0141) **(Figure 5).** LV-EF increased among responders (median change: 0.30%) and declined among non-responders (median change: −2.15%), with a net difference of 2.45% favoring responders during the exercise intervention (p=0.007) (**Table 5**).

**Figure 5.**
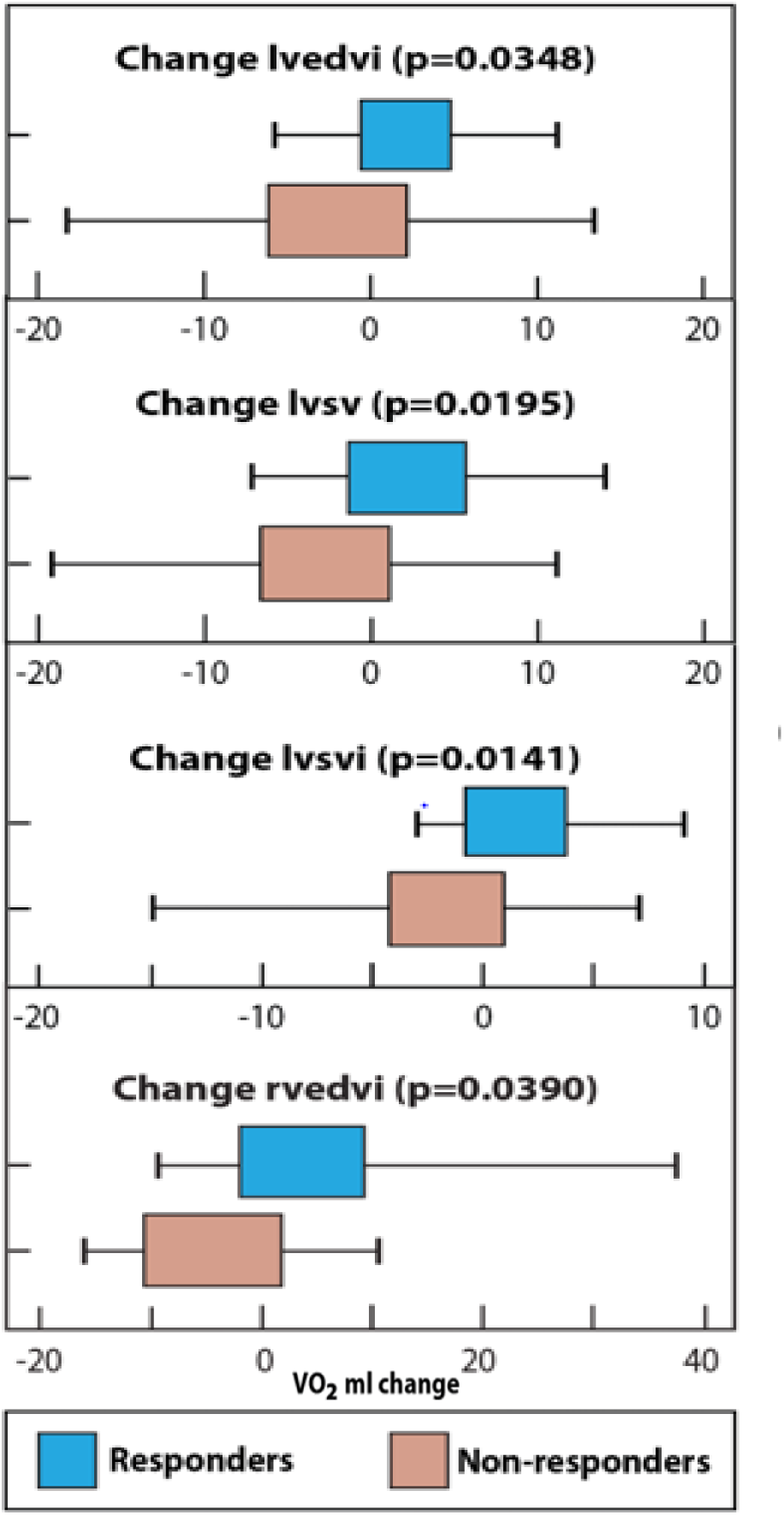
Change in CMR Parameters in Correlation with Differences in VO_2peak_. *Figure demonstrates the significant changes in CMR LV and RV imaging parameters among responders (i.e. ΔVO_2peak_ >1mL/kg/min) and non-responders. Responders demonstrated a change in LV-EDVi (p=0.035), RV-EDVi (p=0.039), and LVSVi (p=0.014) in response to a change in VO_2peak_*.

Polar maps showing segmental circumferential strain from cine tagged acquisition at the short axis basal, mid, and apical levels at baseline (Visit 1) and after the exercise intervention (Visit 2) are shown in **Figure 4**. The color scale indicates peak strain magnitude, with abnormal values in green (≤17%) or yellow (≤10%). High-risk subjects showed improvement in segmental strain, with abnormal myocardial segments decreasing from median 8.9 to 5.9 (≤17%, p=0.014) and from 1.8 to 0.3 (≤10%, p=0.022). Eight of 11 high-risk subjects converted to low-risk at follow-up. Low-risk subjects with predominantly normal segmental strain showed no significant change after the exercise intervention.

Of note, the number of segments with εcc ≤17% correlated with cardiotoxicity risk score (R: 0.29, p=0.040), elevated diastolic and mean blood pressure (R: 0.34, p=0.010), increased LV end systolic volume index (ESVi) (R: 0.34, p=0.010), decreased LV-EF Z-score (R: −0.33, p=0.020), decreased LV mass Z-score (R: −0.3, p=0.030), decreased LV mass/volume Z-score: (R: 0.3, p=0.010), and increased ESFS (R: 0.47, p<0.001). Number of segments with εcc ≤10% correlated with increased LV-ESVi (R: 0.4, p<0.010), decreased LV-EF Z-score (R: 0.46, p<0.001), decreased LV mass/volume Z-score: (R: 0.3, p=0.020), increased ESFS (R: 0.4, p=0.004), decreased RV-EF Z-score (R: 0.39, p=0.007), decreased εcc (R: 0.79, p <0.0001) and ειι (R: 0.49, p<0.0001) and number of segments with εcc ≤17 % (R: 0.56, p<0.0001).

### Predictors of Change in VO_2peak_ at 16 Weeks in Response to Exercise Intervention

Baseline demographic characteristics, CMR parameters and CPET parameters are shown in **Supplemental Table 2**. Responders trended towards a lower LV-EF Z-score, decreased εcc and ειι, and a higher cardiotoxicity risk score (8, IQR 9.5 in responders vs 6.25, IQR 4.5 in non-responders) p=0.025 at baseline. There were no significant differences in baseline CRF between responders and non-responders.

### Relationships Between CRF and CMR Parameters

VO_2peak_ measured after the exercise intervention was positively correlated with expected parameters of physiologic remodeling: end diastolic volume (R: 0.700, p=0.0001), end systolic volume (R: 0.650, p<0.0001), stroke volume (R: 0.756, p=0.0001), and myocardial mass (R: 0.736, p<0.0001).

### Assessment of Physical Activity and Adherence

Fitbit profiles showed a progressive increase in the average number of steps per week during the exercise intervention, peaking at 8 weeks, followed by a decline during the 3rd and 4th months (**Figure 6**).

**Figure 6.**
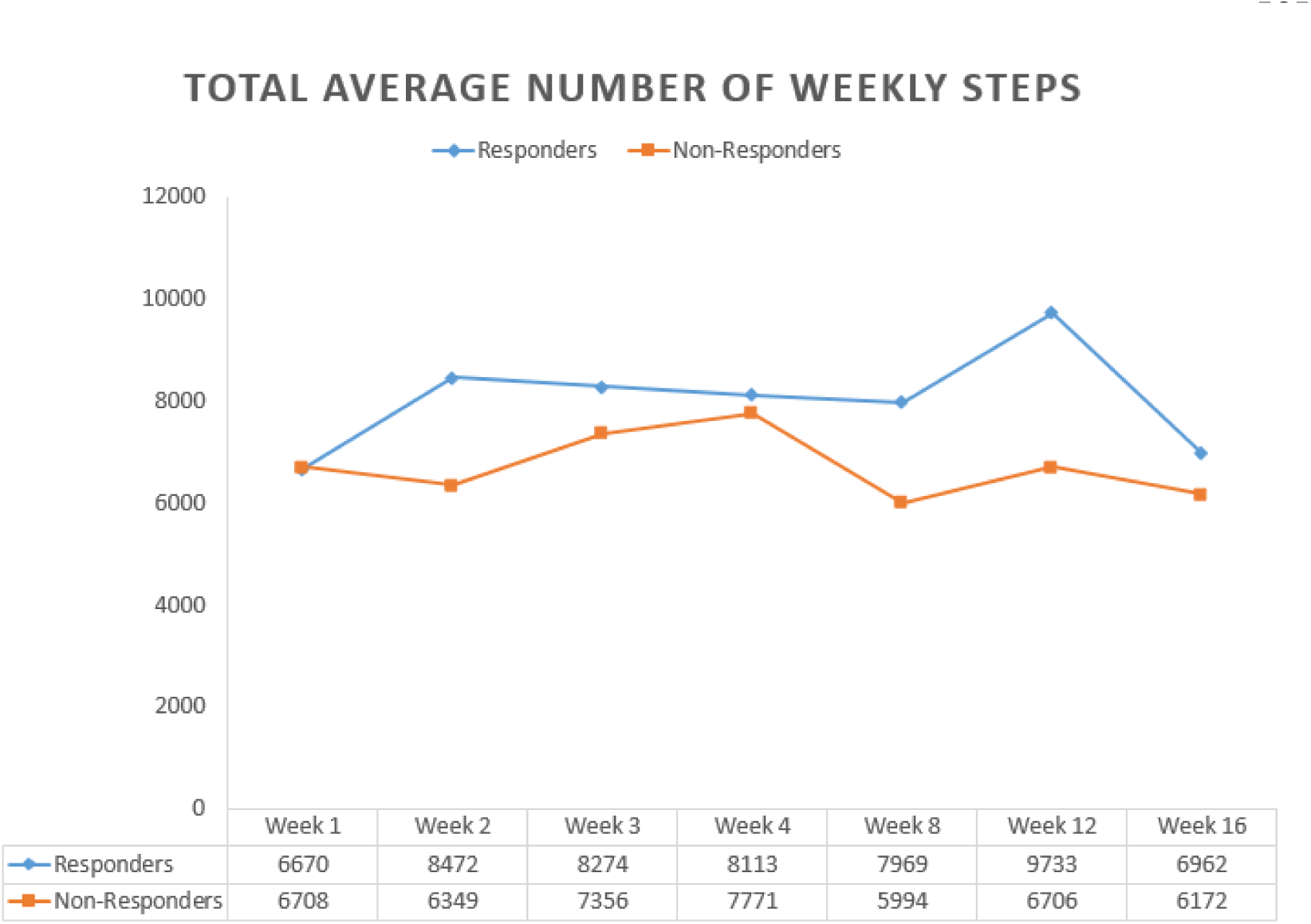
Weekly Average Number of Steps. *The number of steps positively correlated with changes in VO_2peak_ (R: 0.38, p=0.038) and O_2_ pulse (R: 0.3, p=0.004)*.

Adherence was calculated using Fitbit profiles, comparing prescribed versus completed exercise sessions (days with 30-45 minutes of physical activity for 16 weeks). The median adherence was 7 days (IQR 10) of 100% adherence among the 31 subjects. Only 3 subjects had 100% adherence throughout the exercise intervention, whereas 7 subjects did not complete the prescribed sessions in any week. Responders had a higher number of steps starting at week 2, a trend that was consistent throughout the 16-week intervention period (**Figure 6**). The number of steps positively correlated with changes in VO_2peak_ (R: 0.38, p=0.038) and O_2_ pulse (R: 0.3, p=0.004).

## Discussion

This study is the first to evaluate a 16-week supervised exercise intervention as a therapeutic and preventive strategy for CCS. About half of the participants (48.4%) experienced an increase in VO_2peak_ of at least 1 mL/kg/min. Among responders, the median increase was 3.8 mL/kg/min, while non-responders saw a median decline of −1.85 mL/kg/min, resulting in a net difference of 5.65 mL/kg/min. Responders demonstrated physiological cardiac adaptations by CMR, with increased LV-EDVi and RV-EDVi, and improved stroke volume and myocardial segmental strain. EF remained unchanged among responders but declined in non-responders, resulting in a net difference of 2.45% in favor of the responders. Our findings align with previous research that has shown improvements in CRF following exercise in adult and CCS, suggesting a potential cardioprotective effect of aerobic exercise against the cardiotoxic impact of treatment. ^28–30^

Segmental strain dysfunction is a novel CMR biomarker for early identification of patients exposed to anthracyclines who are most at risk for heart failure.^31,32^ CMR strain provides detailed information about the cardiotoxic effects of anthracyclines on myocardial contractility on a regional basis, which allows for better understanding of spatial distribution and temporal progression of regional cardiac dysfunction. Preliminary data in adult CCS post-treatment have demonstrated that segmental strain dysfunction may be a more sensitive parameter to identify patients at risk of heart failure.^22^ In one adult study, the total number of segments expressing ɛcc ≤17% and ɛcc ≤10% identified changes during chemotherapy, allowing for the accurate detection of early onset cardiotoxicity and for strain normalization during implementation of primary and secondary prevention strategies.^32^ This study demonstrates a improvement in segmental myocardial deformation and strain normalization with increased physical activity. The benefits of physical activity were greater in subjects with CVD, as evidenced by the reduction in abnormal myocardial segments in high-risk subjects. ^33^ Previous studies have also demonstrated that exercise attenuates cardiotoxicity as measured by global longitudinal strain.^34^

Previous research has found that CCS exhibit lower V̇O_₂peak_ compared to age-matched non-cancer controls, indicating compromised CRF.^35^ Similarly, this study demonstrated that CCS are deconditioned (mean VO_2peak_ 33.7 ± 9.3 mL/kg/min [range: 16.9 – 57.6]). Functional disability (VO_2peak_ ≤18 mL/kg/min), a strong predictor of adverse long-term outcomes,^36^ was present in four participants. This aligns with previous studies documenting impaired cardiac function and CRF associated with anthracyclines in CCS and women with breast cancer.^35,37^

In youth, VO_2peak_ predicts various health indicators including cardio-metabolic health, premature CVD, academic achievement, and mental health.^23,38,39^ Exercise training is widely accepted as a preventive and therapeutic strategy for various CVD.^38,40^ Additionally, it is now an accepted component of the therapeutic regimen for patients with heart failure.^41,42^ A systematic review highlighted that exercise interventions are both feasible and effective in promoting physical activity among CCS. These interventions not only enhance physical fitness but also reduce adverse late effects associated with cancer treatments.^43^

Research on exercise interventions in CCS has identified several challenges: maintaining high motivation, engaging subjects with a negative attitude towards physical activity, and achieving long-term participation.^44^ In adults with cancer, a wide variety of exercise modes have been studied including walking, cycling, strength training, and yoga. Among 28 studies reviewed in a Cochrane database, 13 included multiple modes of exercise.^45,46^ As demonstrated by this pilot study, a supervised exercise intervention is feasible in CCS. The main barrier to participation in our cohort was access to a local YMCA. Only 7 participants did not complete the prescribed number of exercise sessions in any week throughout the study time period. Customizing exercise interventions to individual patients can overcome participation challenges. Personalized exercise interventions are better received as subjects have a diverse level of activity requirements and preferences.^47^ The customized exercise intervention based on the subject’s preferred exercise modality was well received.

The exercise intervention in CCS showed significant individual variation in CRF response and adherence, highlighting the need for a targeted, individualized approach. Barriers included access to a local YMCA and high trainer turnover, leading to inconsistent communication. These findings underscore the importance of implementing targeted interventions to address deconditioning in childhood cancer survivors, aiming to improve their overall health and quality of life. Notably, mobile health strategies have emerged as promising tools to encourage physical activity in this population and lay the foundation for the use of a theory-based mobile application designed to encourage sustainable health behavior changes, reduce sedentary habits, and improve compliance and adherence to the exercise intervention. ^48^

### Study limitations

This pilot study was designed to assess feasibility of a customized exercise intervention and impact on CMR imaging parameters with a need for a larger randomized control study to confirm the results. Although our study design did not include a control group, non-responders, who were less adherent and completed fewer steps as demonstrated by the Fitbit output, serve this purpose.

## Conclusions

Exercise intervention in CCS offers a non-pharmacological mechanism to favorably modulate physiologic cardiac remodeling in CCS, and CMR offers mechanistic end-points that can be measured as a result of the intervention. Knowledge gained from this study will provide insight into behavioral strategies through the use of mobile technology to motivate adoption of healthier lifestyle behaviors such as increases in physical activity and improved cardiovascular health in CCS.

## Funding

This study was supported by the Maximilian E. & Marion O. Hoffman Foundation

## Disclosures

The authors declare that they have no competing interests.

## Data Availability

All data supporting the findings of this manuscript are available from the corresponding author upon reasonable request.

## Acknowledgments

The authors thank James Whayne, MSc, and Farouk Osman, BS, for their assistance in analysis and figure preparation on segmental strain. We also appreciate the support of the research teams at Connecticut Children’s and Nationwide Children’s Hospital. We acknowledge funding from The Heart Center at Nationwide Children’s Hospital Intramural Fund Mechanism and thank the regional YMCAs for their exercise programs, which made this study feasible.

## List of Abbreviations

CCS: childhood cancer survivors
CVD: cardiovascular disease
CRF: cardiorespiratory fitness
VO_2peak_: peak oxygen consumption
LV: left ventricular
CMR: cardiac magnetic resonance imaging
CPET: cardiopulmonary graded exercise stress test
RV: right ventricular
SV: stroke volume
EF: ejection fraction
ESFS: end-systolic fiber stress
Ειι: peak longitudinal strain magnitude
ɛcc: peak circumferential strain magnitude
EDVi: end diastolic volume index
ESVi: end systolic volume index

---

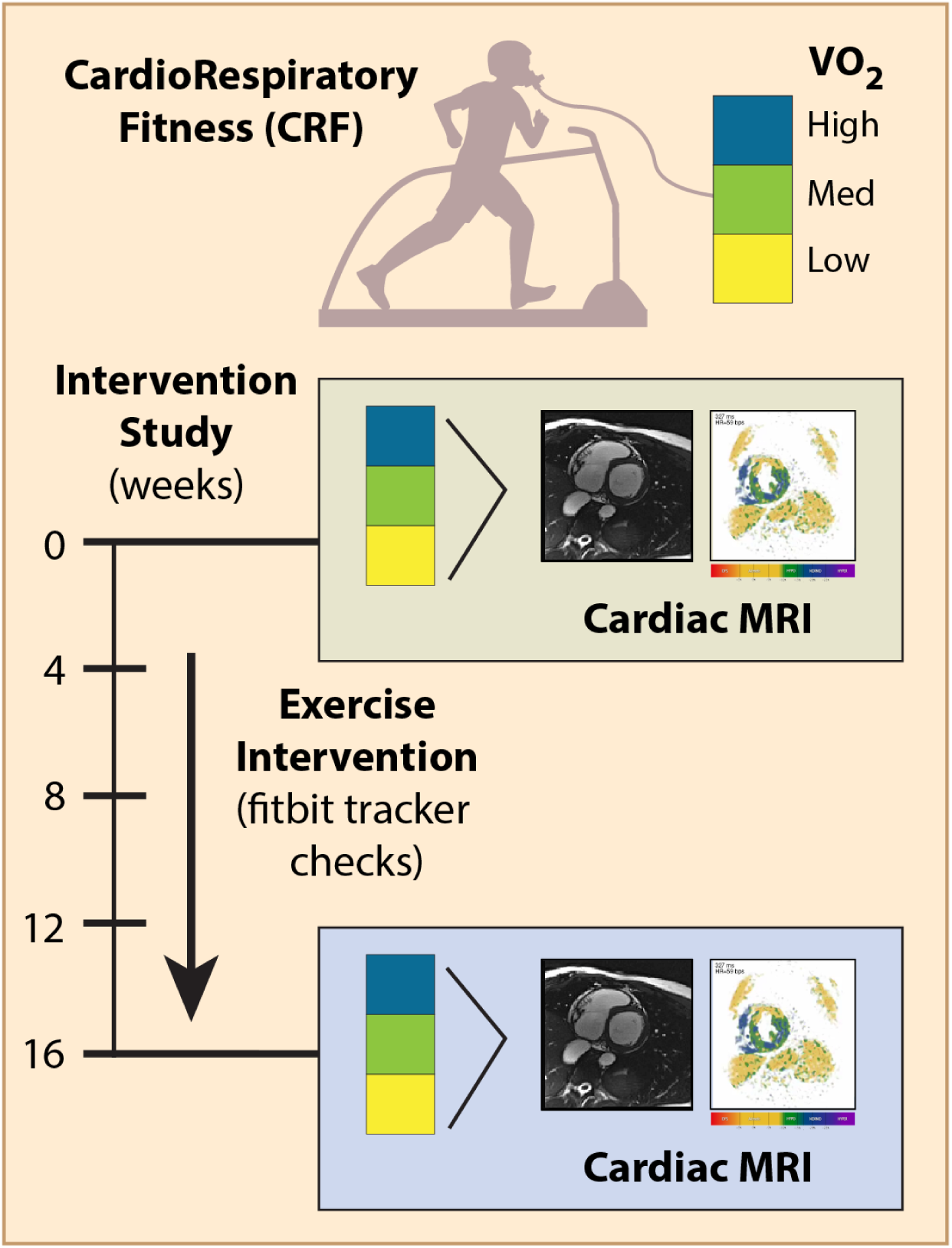
Exercise Intervention Protocol [CENTRAL ILLUSTRATION]. *In this 16-week supervised exercise intervention, we used local YMCA trainers who were trained in the Livestrong program and comfortable working with CCS. Patients chose from a set of aerobic and resistance exercise modalities. Participants underwent cardiotoxicity risk stratification, a CPET, and CMR imaging at baseline and following the 16-week exercise intervention (15 to 45 minutes, three to five days a week, at 50-80% of their age adjusted maximum heart rate as determined by CPET)*.

## Notes

### Competing Interest Statement

The authors have declared no competing interest.

### Clinical Trial

Trial registration: NCT, NCT04036032. Unique Protocol ID: 14-110. Registered 25 July 2019 - Retrospectively registered, https://classic.clinicaltrials.gov/ct2/show/NCT04036032

### Author Declarations

This study was a prospective multicenter pilot study. This study was approved by the Institutional Review Board at Connecticut Children's and Nationwide Children's Hospital.

